# Nutrition-sensitive Agriculture – Is it possible in Morogoro Rural District? Awareness and practices among farmers

**DOI:** 10.1101/2022.06.29.22276899

**Authors:** Innocent Sanga, John Msuya

**Affiliations:** Department of Human Nutrition and Consumer Sciences, College of Agriculture, Sokoine University of Agriculture, P.O. Box 3006, Chuo Kikuu, Morogoro, Tanzania

**Keywords:** nutrition-sensitive agriculture, nutrient dense foods, home gardening, livestock keeping, biofortified crops

## Abstract

According to FAO (2017), nutrition-sensitive agriculture is an approach that seeks to ensure the production of a variety of affordable, nutritious, culturally appropriate and safe foods in adequate quantity and quality to meet the dietary requirements of populations in a sustainable manner. This study focused on analyzing ways in which the concept is implemented and assessing its awareness among the key nutrition stakeholders. These included government officials from village, ward and district levels, NGOs, agriculture inputs suppliers, food vendors and grain millers. Cross - sectional study design was employed and information from respondents were collected by using semi-structured questionnaires, key informant interviews and transect walk. Data analysis was conducted by using SPSS version 20. It was found that home gardening (85.3%) and livestock keeping (57.4%) were the most practiced ways of nutrition-sensitive agriculture. Other ways, including use of biofortified crops, fish farming, use of watering techniques in production of fruits and vegetables, use of soil fertility conserving techniques and using improved food crop varieties were hardly practiced. Above 90% of household heads knew the importance of agriculture to nutrition being a source of food for household consumption, a source of income and employment, makes easy to access nutrient dense foods in the household and ensuring food security. The study also observed high awareness about the concept even though, the actual implementation was low. It was concluded that, low economic status among the rural poor farmers is the major reasons that have led to the observed low practices of other studied ways of agriculture that are nutrition sensitive. It is recommended that, all key stakeholders should work together to subsidize seeds and other agro-inputs which are needed by the farmers to practice these ways.

## Background

Nutrition-sensitive Agriculture is an approach that seeks to ensure the production of a variety of affordable, nutritious, culturally appropriate and safe foods in adequate quantity and quality to meet the dietary requirements of populations in a sustainable manner (FAO, 2017). The concept has recently been a key focus area by a number of governments, donor agencies, and development organizations including FAO, IFAD and IFPRI (Chipilli and Msuya, 2016; FAO, 2017; Ruel *et al*., 2018). Agriculture employs majority of poor population in most of the developing countries who are in turn vulnerable to malnutrition (Gillespie *et al*., 2015). Tanzania is faced with high malnutrition prevalence rates where-by 34% children under five years of age were stunted and 58% of women of reproductive age were anemic (URT, 2016a). Malnutrition is more prevalent in rural areas where majority of the residents earn their living through agricultural production (Bundala *et al*., 2019). For example, stunting affects children under five years of age more in rural areas, at 38%, than urban areas, at 25%; (World Bank, 2013; URT, 2016a). Similar trends have been observed in the recent Tanzania national nutrition survey of 2018 (URT, 2018). Malnutrition in Morogoro region has remained to be a huge problem. Current data from the Tanzania National Nutrition Survey (TNNS) indicates that 26.4 % of children underfive years of age are stunted, 12.1% are underweight and 6% are wasted (URT, 2018).

Agriculture becomes nutrition-sensitive when all activities in the whole food value chain from production to consumption incorporate nutrition objectives (URT, 2011; Gillespie *et al*., 2015; Herforth, 2013;). The production stage in the agro-food system provides an easy stage through which the household can produce and ensure availability and consumption of nutritious food items. Biofortification of staple crops, home gardening for nutrient-dense vegetables and fruits, fish farming, livestock keeping and nutrition-sensitive agriculture production system including irrigation programs and agriculture extension services are ways of ensuring Nutrition-sensitive agriculture increase access to nutritious foods of both crop and animal origin in households (Shetty, 2018; Ruel *et al*., 2018).

In that respect, it is critically important that all key actors or stakeholders, are well informed, or aware about the concept and benefits of practicing the various techniques of agriculture that is nutrition sensitive. These actors include food producers (basically farmers), food processors and traders, caregivers and ultimately consumers themselves.

This study intends to assess the awareness and the ways in which the ways of nutrition-sensitive agriculture are practiced by farmers in the food value chain in Morogoro rural district in Tanzania.

## Materials and methods

The study was conducted in Morogoro rural district in Morogoro Region, Tanzania. The district is divided into 6 divisions namely Bwakira, Mvuha, Mikese, Mkuyuni, Matombo, Ngerengere. With a total of 31 wards including Mkuyuni and Mvuha each having 28 and 27 villages respectively. Moreover, the district has 3 agro-ecological zones, savannah, low mountainous and mountainous zones, where 82% of the adult people earns their livelihood from agriculture (NBS, 2012).

Major economic activities through which people earn their living includes agriculture, forestry, fishery, beekeeping, mining and other businesses. Main food crops grown by majority of the farmers includes staple crops such as maize, paddy, beans and sorghum while chicken, cattle and goats are the major livestock domesticated by livestock keeping households. According to Morogoro district profile, the district is faced with malaria, pneumonia and acute respiratory infections are some of the major health problems (URT, 2016b).

Cross - sectional study design was used in which data collection was done at one point in time. Morogoro Rural District was purposively selected from Morogoro Region because of high likelihood of households practicing agriculture, both crop farming as well as livestock keeping.

Two wards from the District namely, Mvuha and Mkuyuni were purposively selected because of differences in agro-ecological conditions. Whereby Mvuha has a relatively flatland topography and Mkuyuni is mountaneous in nature. Random sampling was used to obtain the households that practice agriculture either through farming/crops cultivation or livestock keeping. Villages were purposively sampled, two of them were located around the ward center while one was located far from center of the ward. Mkuyuni, Kibwaya and Changa villages were selected from Mkuyuni Ward, with Changa being located far from the ward centre while those from Mvuha ward included Dalla, Mvuha and Tulo. The latter being located relatively far from the ward centre.

Households were then randomly selected from the list of all households in each village while actors from sectors key to nutrition were from several village, ward and district departments. The techniques of nutrition sensitive agriculture which were studied included fish farming, sustaining production of nutrient dense crops through applying soil fertility conserving techniques and minimizing the use of industrial fertilizer through the use of farm yard manures. Livestock keeping (include small livestock), watering techniques in production of fruits and vegetables, use of improved food crop varieties, home gardening and biofortified food crops were other ways that were studied.

Also awareness of about nutrition sensitive agriculture was studied by considering 7 aspects; ensuring food security, source of employment, enabling income gain making the purchase food easier, allowing for flexibility of women to care for their children in the household source of food for household consumption,, allows for gender equity in the household, improving dietary diversity in the household/consumption of various food groups and making easy to access nutrient dense foods in the household

Household heads and stakeholders from sectors key to nutrition were the two categories of respondents involved in the study. The sample size was computed according to Kothari (2004) obtaining 226 households but due to economic constraints total of 115 households representing household heads were recruited in the study, 57 and 58 household heads in mkuyuni and mvuha wards respectively. Other stakeholders included government officials from district, ward and village levels. Others (30 of them) were from NGOs, food vendors, agriculture inputs suppliers and grain millers.

Data were collected by interviewing the head of households using a semi-structured questionnaire, while the key informants were interviewed using checklist questions formulated accordingly. Participatory transect walks and observation were used to complement information from the interview data. More illustration is given by Sanga and Msuya (2022).

Quantitative data from household interviews were analyzed by using Statistical Product and Service Solutions version 20. Frequencies and percentages for studied aspects of practicing nutrition-sensitive agriculture and awareness parameters were calculated from descriptive statistics. Overall, statistical tests were 2-tailed, and P values ≤ 0.05 were considered statistically significant. Qualitative data were summarized into key themes and reported as said. Information from transect walks were summarized and presented in a transect sketch.

## Results

### Households characteristics

Characteristics considered included demography, ethnicity, economic activities, land ownership and main source of food. These were categorized with respect sex of the household head. Results shows that there were more male in all the characteristics of the sampled households than were females. This is because there were more male than female headed households with majority being middle aged adults among both sex categories. There were more primary educated individuals with more males (58.3%) than females (18.3%). On the other hand, results indicate that many household heads were married, with more males (71.3%) than females (9.6%). Also, majority of them originated from the study areas, with just a few of them who moved in from other areas. Furthermore, the results indicate that many households owned small land plots between 0.5 and 2 acres while in all categories of household food source, results indicate a difference in the wild foods as source of food in the household with more males (51.3%) than female headed households (20%) confirming to utilize them. Lastly, among the studied economic activities, apart from farming which was practiced by all households regardless of the sex of the household head, other activities highlighted a difference with more males than females dominating each category.

### Ways Nutrition - Sensitive Agriculture is implemented

Table 2 shows that home gardening was the most practiced way followed by livestock keeping. Other less important ways included the use of biofortified food crops, the use of improved food crop varieties, sustaining production of nutrient dense crops through applying soil fertility conserving techniques, watering techniques and minimizing the use of industrial fertilizers through the use of farm yard manures in production of food crops and fish farming. The production of biofortified food crops was the only way that showed significant difference to be practiced in the study area. No significant difference was observed for other studied ways.

**Table 1:**
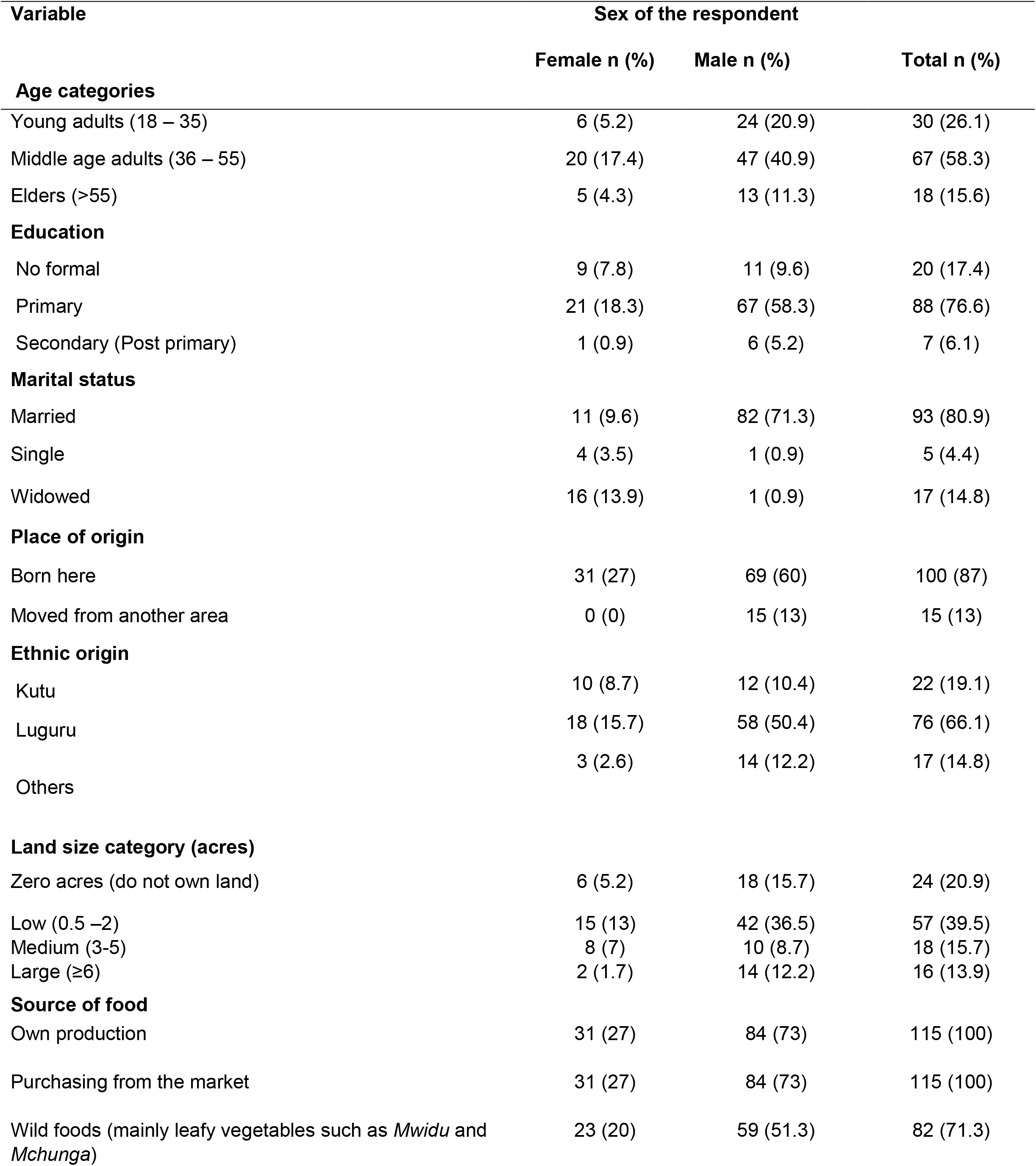
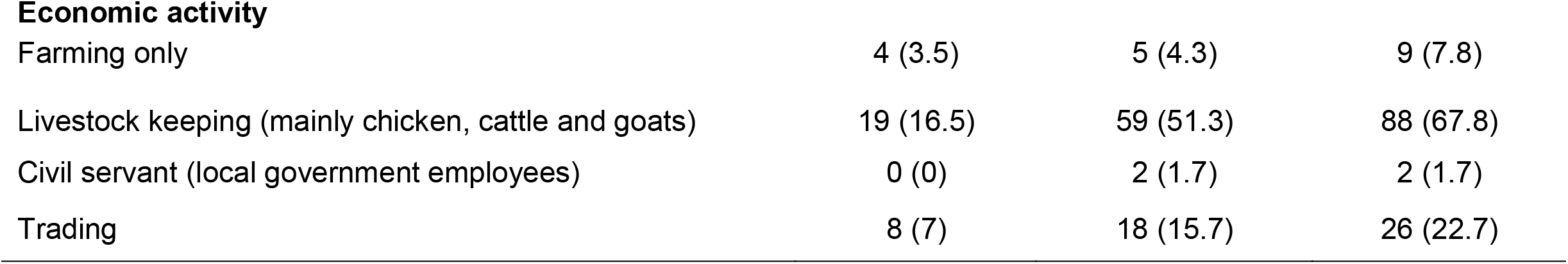
Characteristics of household heads according their sex.

| Variable | Sex of the respondent |  |  |
| --- | --- | --- | --- |
|  | Female n (%) | Male n (%) | Total n (%) |
| **Age categories** |  |  |  |
| Young adults (18 – 35) | 6 (5.2) | 24 (20.9) | 30 (26.1) |
| Middle age adults (36 – 55) | 20 (17.4) | 47 (40.9) | 67 (58.3) |
| Elders (>55) | 5 (4.3) | 13 (11.3) | 18 (15.6) |
| **Education** |  |  |  |
| No formal | 9 (7.8) | 11 (9.6) | 20 (17.4) |
| Primary | 21 (18.3) | 67 (58.3) | 88 (76.6) |
| Secondary (Post primary) | 1 (0.9) | 6 (5.2) | 7 (6.1) |
| **Marital status** |  |  |  |
| Married | 11 (9.6) | 82 (71.3) | 93 (80.9) |
| Single | 4 (3.5) | 1 (0.9) | 5 (4.4) |
| Widowed | 16 (13.9) | 1 (0.9) | 17 (14.8) |
| **Place of origin** |  |  |  |
| Born here | 31 (27) | 69 (60) | 100 (87) |
| Moved from another area | 0 (0) | 15 (13) | 15 (13) |
| **Ethnic origin** |  |  |  |
| Kutu | 10 (8.7) | 12 (10.4) | 22 (19.1) |
| Luguru | 18 (15.7) | 58 (50.4) | 76 (66.1) |
| Others | 3 (2.6) | 14 (12.2) | 17 (14.8) |
| **Land size category (acres)** |  |  |  |
| Zero acres (do not own land) | 6 (5.2) | 18 (15.7) | 24 (20.9) |
| Low (0.5 –2) | 15 (13) | 42 (36.5) | 57 (39.5) |
| Medium (3-5) | 8 (7) | 10 (8.7) | 18 (15.7) |
| Large (≥6) | 2 (1.7) | 14 (12.2) | 16 (13.9) |
| **Source of food** |  |  |  |
| Own production | 31 (27) | 84 (73) | 115 (100) |
| Purchasing from the market | 31 (27) | 84 (73) | 115 (100) |
| Wild foods (mainly leafy vegetables such as *Mwidu* and *Mchungu*) | 23 (20) | 59 (51.3) | 82 (71.3) |
| **Economic activity** |  |  |  |
| Farming only | 4 (3.5) | 5 (4.3) | 9 (7.8) |
| Livestock keeping (mainly chicken, cattle and goats) | 19 (16.5) | 59 (51.3) | 88 (67.8) |
| Civil servant (local government employees) | 0 (0) | 2 (1.7) | 2 (1.7) |
| Trading | 8 (7) | 18 (15.7) | 26 (22.7) |

**Table 2:**
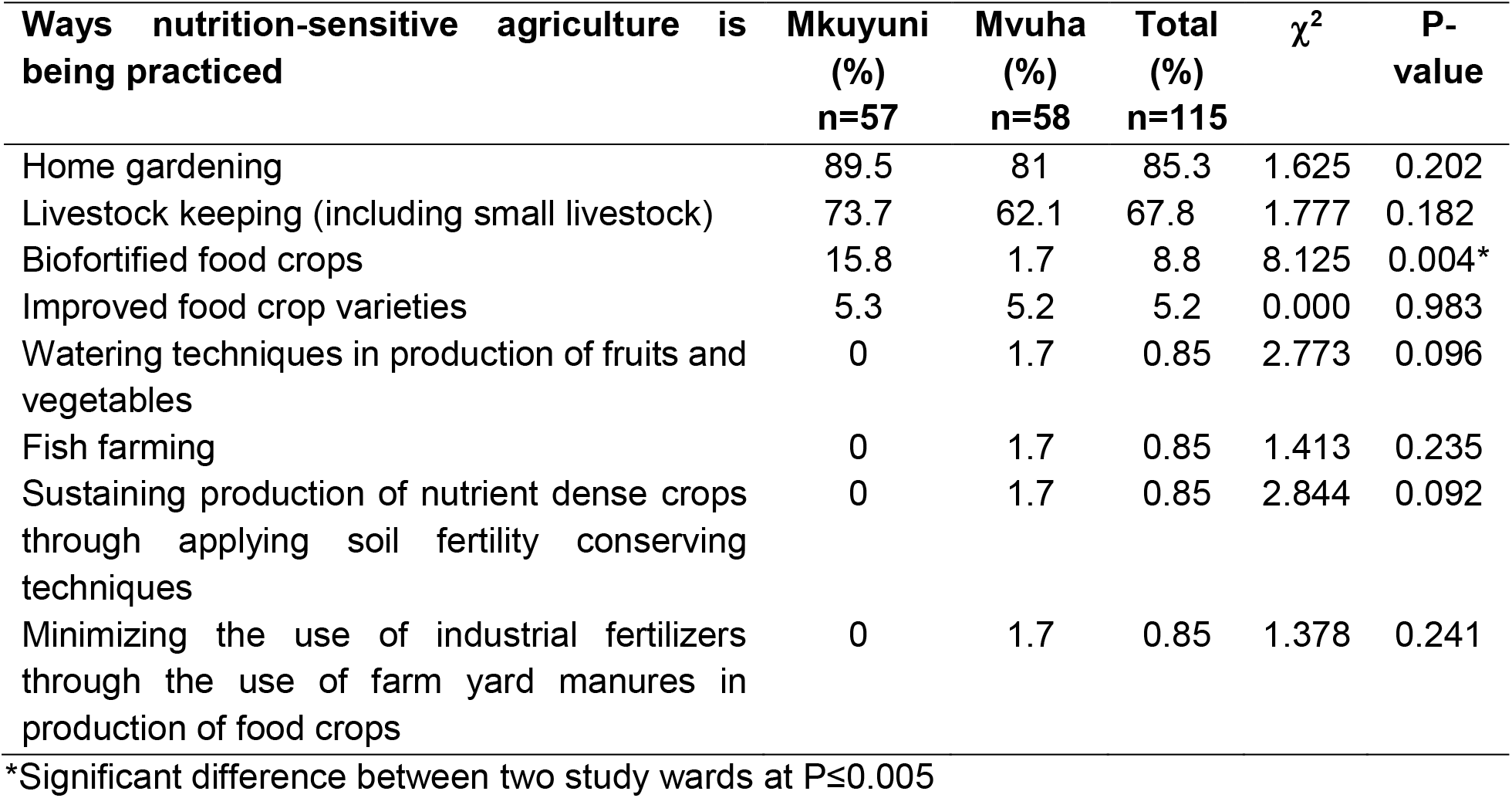
Households practicing ways of nutrition-sensitive Agriculture in the study wards.

| Ways nutrition-sensitive agriculture is being practiced | Mkuyuni (%)<br>n=57 | Mvuha (%)<br>n=58 | Total (%)<br>n=115 | $\chi^2$ | P-value |
| --- | --- | --- | --- | --- | --- |
| Home gardening | 89.5 | 81 | 85.3 | 1.625 | 0.202 |
| Livestock keeping (including small livestock) | 73.7 | 62.1 | 67.8 | 1.777 | 0.182 |
| Biofortified food crops | 15.8 | 1.7 | 8.8 | 8.125 | 0.004* |
| Improved food crop varieties | 5.3 | 5.2 | 5.2 | 0.000 | 0.983 |
| Watering techniques in production of fruits and vegetables | 0 | 1.7 | 0.85 | 2.773 | 0.096 |
| Fish farming | 0 | 1.7 | 0.85 | 1.413 | 0.235 |
| Sustaining production of nutrient dense crops through applying soil fertility conserving techniques | 0 | 1.7 | 0.85 | 2.844 | 0.092 |
| Minimizing the use of industrial fertilizers through the use of farm yard manures in production of food crops | 0 | 1.7 | 0.85 | 1.378 | 0.241 |

\*Significant difference between two study wards at $P \leq 0.005$

Agriculture officers from the district and ward levels reported that it is not surprising that people in the area do practice home gardening more because there has been more emphasis of the practice community members to grow vegetables. The use of biological/organic techniques for control of crop pests and diseases, other simple and useful agricultural practices such as mulching for preserving soil moisture have been introduced. Farmers are also encouraged to use improved crop varieties and organic manure in place of inorganic industrial made fertilizers.

However, some ward agriculture officers, were concerned that adoption of growing nutrient-dense crops such as yellow maize and Orange Fleshed Sweet Potatoes was still very low blaming the cause to be low attendance of the farmers during the training gatherings. On the other hand, it was noted by the livestock officers that people in this area avoid keeping large animals such as cattle and goats because they fear the activity is too labor intensive. They blamed that to be one of major reasons for low availability of animal source foods, especially meat and milk in the area. Same opinion was also reflected by the District Nutrition Officer. Cattle were only kept by few migrants of Masai and Sukuma ethnic origin.

From the transect sketch (Sanga & Msuya, 2022), home gardens of vegetables and fruits were established around homes and others in small farm plots. Livestock keeping was practiced by very few, and majority kept poultry birds mainly chicken, while a very few of them kept other livestock such as goats and cattle. It was also found that few individuals practiced watering techniques in production of food crops except those individuals who grew crops along river banks. It was observed that although Mkuyuni ward was mountainous with steep slopes, very few individuals in the ward were practicing contour farming and zero tillage to reduce soil erosion. Over all in the two wards, very few farmers were growing biofortified crop varieties.

### Perceived liking for the different aspects of nutrition-sensitive agriculture

Study subjects were asked to indicate their liking of the different aspects of the concept using five-point ranking scale (with dislike completely scoring the lowest i.e 1 and likes the most scoring the highest i.e 5). This was done irrespective of whether the respondent was doing it or not.

Table 3 indicate the mean average scores for each tested aspect. Home gardening scored the highest whereas other ways followed in scores. The use of farm yard manures in production of food crops, sustaining production of nutrient-dense crops through applying soil fertility conserving techniques, livestock keeping, improved food crop varieties and minimizing the use of industrial fertilizers through the use of farm yard manures in production of food crops, had the least mean. Other aspects scored lower mean values included watering techniques in production of fruits and vegetables, fish farming and production of biofortified food crops.

**Table 3:** Mean scores on the tested aspects of nutrition-sensitive agriculture.

| **Aspects of nutrition-sensitive agriculture** | **Mean score**<br>**(Mean ± SD)** |
| --- | --- |
| Home gardening | 4.36 ± 0.752 |
| Production of biofortified food crop varieties | 3.66 ± 0.724 |
| Livestock keeping (including small livestock) | 4.05 ± 0.560 |
| Improved food crop varieties | 4.03 ± 0.561 |
| Watering techniques in production of fruits and vegetables | 3.56 ± 0.774 |
| Sustaining production of nutrient dense crops through applying soil fertility conserving techniques | 4.20 ± 0.610 |
| Minimizing the use of industrial fertilizers through the use of farm yard manures in production of food crops | 4.00 ± 0.662 |
| Fish farming | 3.20 ± 0.840 |

Figure 1 shows ranking of studied ways. Home gardening was liked the most by majority of the respondents whereas other ways such as sustaining production of nutrient dense crops through applying soil fertility conserving techniques, minimizing the use of industrial fertilizers through the use of farm yard manures in production of food crops, livestock keeping, use of improved food crop varieties, production of biofortified food crops, watering techniques in production of fruits and vegetables and fish farming followed. Very few individuals indicated neutral or dislike on the studied ways indicating low engagement or readiness to practice a particular way.

**Figure 1:**
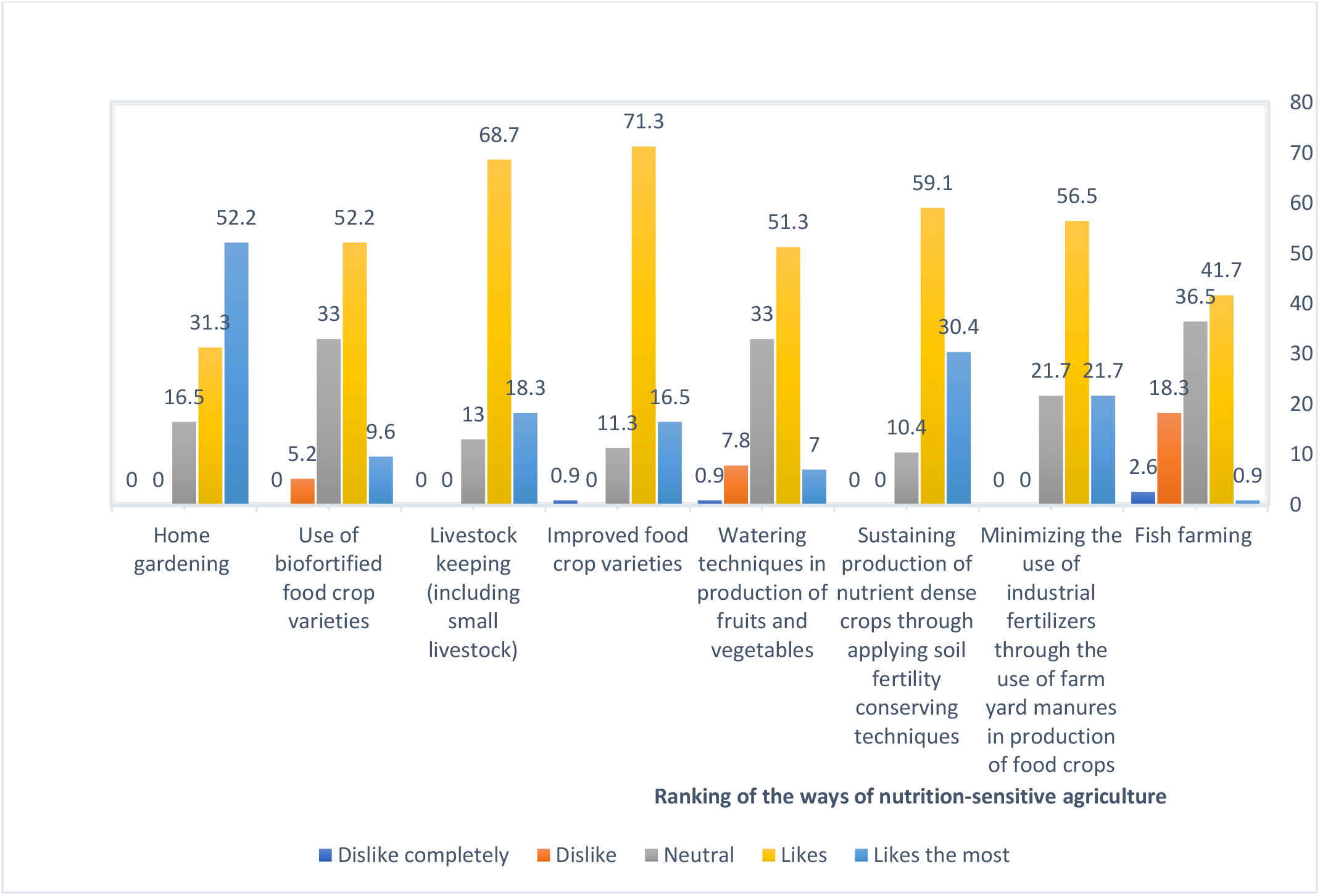
Ranking preference of the studied ways of nutrition-sensitive agriculture.

### Awareness about nutrition–sensitive agriculture

In assessing the awareness of the farmers about nutrition-sensitive agriculture, two issues were considered. Firstly, was awareness of the household heads on how agriculture can benefit nutrition, and secondly, the extent to which nutrient-dense foods are consumed by the respondents.

#### (i) Awareness on how agriculture can benefit nutrition

Eight aspects were used to assess household head awareness through which agriculture can benefit nutrition. The eight aspects were the following:

▪ Source of employment
▪ Ensuring food availability
▪ Improving dietary diversity in the household/consumption of various food groups
▪ Makes easy to access nutrient dense foods in the household
▪ It allows for flexibility of women to care for their children in the household
▪ Enabling income gain therefore making easy to purchase food
▪ Source of food for household consumption
▪ Allows for gender equity in the household

As indicated in table 4, it was found that, majority of the study subjects were aware about the importance of agriculture to nutrition where as low awareness were observed in just one of the studied benefits.

**Table 4:** Respondents awareness on how agriculture can benefit nutrition.

| **Benefit** | **Yes**<br>**n (%)** | **No**<br>**n (%)** |
| --- | --- | --- |
| Source of employment | 115 (100) | 0 |
| Enable income gain therefore making easier to purchase food | 114 (99.1) | 1 (0.9) |
| Source of food for household consumption | 115 (100) | 0 |
| Allows for gender equity in the household | 82 (71.3) | 33 (28.7) |
| Makes easy to access nutrient dense foods in the household | 112 (97.4) | 3 (2.6) |
| Ensures food security | 115 (100) | 0 |
| Improve dietary diversity in the household/consumption of various food groups | 94 (81.7) | 21 (18.3) |
| It allows for flexibility of women to care for their children in the household | 67 (58.3) | 48 (41.7) |

The awareness scores on the eight aspects of how agriculture can benefit nutrition was placed into three categories. The highest score was 8 (aware about all the 8 aspects and the minimum was zero (not knowing any of them). three categories were formulated from the scores as low, medium and high awareness whereby the results are shown in table 5.

**Table 5:**
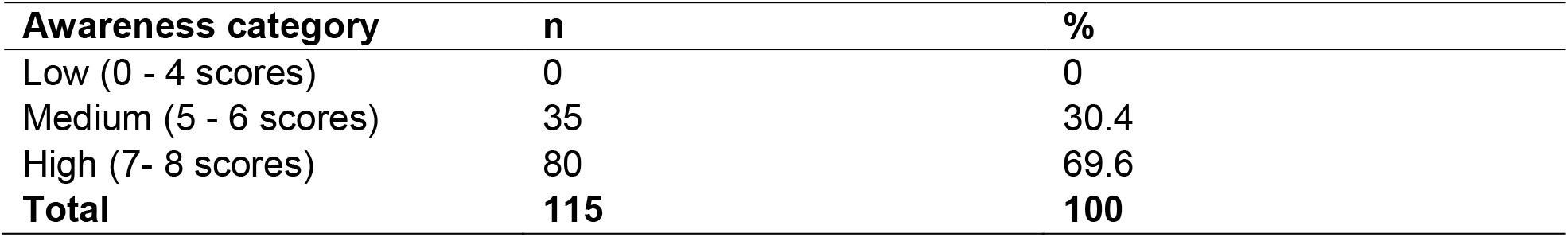
Category of awareness based on the respondents scores of eight aspects that were assessed.

#### (ii) Consumption of nutrient-dense foods in the household

Results about the consumption of nutrient-dense foods is shown in figure 2. Vegetables had the highest consumption rate while medium consumption was observed for legumes and fruits. Foods of animal source were consumed by very few individuals and in that respect, good proportions had never consumed them.

**Figure 2:**
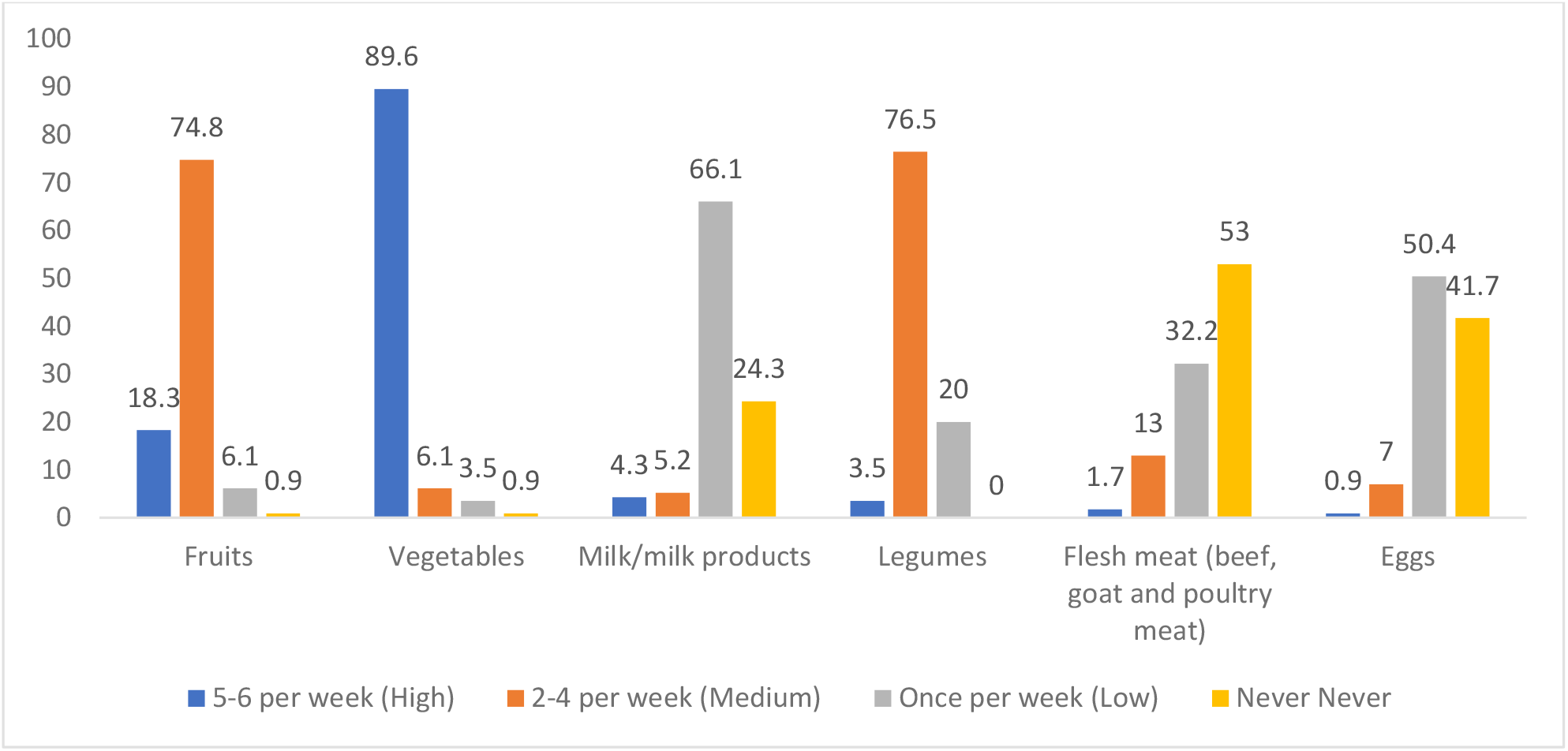
Percentage showing frequency of consumption of different food groups.

## Discussion

### Ways of nutrition-sensitive agriculture implemented in the study area

The focus of the current study has been mostly on the food production and to a small extent the consumption step on food chain. Selected ways in which nutrition–sensitive agriculture can be implemented were assessed. They included livestock keeping for obtaining foods of animal origin, home gardening, use of biofortified food crop varieties, improved food crop varieties, watering techniques, fish farming, sustaining production of nutrient dense crops through applying soil fertility conserving techniques and minimizing the use of industrial fertilizers through the use of farm yard manures in production of food crops. Each one of these different ways are discussed here separately.

#### (i) Home gardening

One of the most convenient way of ensuring food availability and access in the household is through home gardening (Mustafa *et al*., 2021), especially for fruits and vegetables. The practice minimizes the cost of purchasing these foods from the markets, and therefore save the money to obtain other food varieties which diversify the household diet. Majority of the study subjects reported to practice home gardening, which was also reflected by their high rate of consuming vegetables and fruits (as shown in figure 2). Weinberger (2013) reported similar results that home gardens contributed to more diversified diets through consumption of nutritionally-rich foods including fruits and vegetables. The study also found that home gardening was practiced as a common task around homes.

Unfortunately, the interviewed agriculture extension officers at the ward level indicated that growing of home gardens was not part of the district agricultural plan implying that it was not emphasized compared to the staple crops such as maize and paddy. It is not surprising that very little technical support was given to the farmers on growing vegetables and fruits in home gardens. Nordhagen *et al*. (2019) suggested that, if appropriate technical support to the farmers is provided, production and farmers income which were low in the study area could be maximized.

Results show that very few respondents practiced watering in production of fruits or vegetables. A few were engaged in small scale production of vegetables along the water streams as revealed by the transect walks (Sanga and Msuya, 2022), and individual farmers reported that they use these water sources to irrigate their small plots. This observation could be due to issues concerning availability of reliable water supply systems and ease with which they have access to them as it was observed that there were poorly watering schemes. This has also been indicated in the Tanzania agriculture policy as one of the challenges facing the agriculture sector (URT, 2013). Passarelli *et al*. (2018) reported that, small scale irrigation/watering schemes are viewed as less desirable and inefficient in many parts of Tanzania. They also showed that most of the traditional and informal schemes are considered illegal given that farmers do not have formal water user rights. Drechsel et al. (2006) also noted that in most of the developing countries, informal watering systems receive little recognition but they are highly used by most of the small holder farmers.

When watering schemes are developed and advanced, it is possible to improve agricultural productivity. It has been reported that using buckets and watering cans takes more than 2500 hours per year to irrigate one hectare of vegetables in Tanzania whereas approximately 250 hours are spent for the same task using motorized pumps (Keraita and de Fraiture, 2011). For activities such as home gardening which are normally controlled by women, the use of mechanized irrigation can be an important entry point for women’s empowerment, by saving time and a good source of household income (Malapit *et al*. 2013; Domènech, 2013; Sraboni *et al*. 2014; Domènech, L. & Ringler. 2013; Farnworth *et al*. 2016).

#### (ii) Livestock keeping

Keeping of livestock around home enables availability of protein and micronutrient rich food sources which are important for health of household members especially for the children (FAO, 2017). This study found that chicken were the main livestock kept in households. Similar findings were reported by De Bruyn *et al*. (2018), Bundala *et al*. (2019) and Bellows *et al*. (2020) in other parts of Tanzania. It was common that chicken were left to move around freely in search of their feeds (i.e. free range). The method of keeping poultry birds was convenient for many poor rural households who may not be able to afford the costs of feeding the birds when confined. It has been reported that, convenience of keeping the animals tends to drive the owner to keep certain species of livestock than others (Matthiesen *et al*. 2011).

Ethnicity of the study subjects as one of the social cultural factors influencing agricultural production habits of the people, was observed to be associated with the type of livestock the household kept (Hetherington *et al*., 2017). It was observed that individuals who migrated into the study area, mainly Maasai and Sukuma people were the ones involved in keeping large animals such as cattle and goats as compared to the natives who mainly kept small animals especially poultry such as chicken and ducks (URT, 2016b). This is because keeping of these livestock types is their lifestyle and main economic activity. Bundala *et al*. (2019) also found similar patterns of livestock keeping in rural parts of Tanzania. It was reported that fear of cattle raiding by pastoralist people (eg. Maasai) was also hindering the natives from attempting to keep large animals, especially cattle, while Muslim beliefs were discouraging the keeping of swine or pigs.

#### (iii) Use of biofortified food crops

According to FAO (2017), in order to increase the levels of micronutrient intakes, biofortification can be used to achieve this goal especially when the biofortified food crop is a staple food in the target population. Stein *et al*. (2007) and Mustafa *et al*. (2021) reported that, as one of the plant breeding techniques that can enhance micronutrients status, biofortification has been reported to be cost effective way of providing high nutrient dense staples. Its limitation is that, the approach alone cannot be relied to treat the severe cases of micronutrients deficiencies especially among women and children who have higher demands but it can serve to provide daily dose of micronutrients to help prevent the deficiencies throughout their lives (Ruel *et al*., 2013; Shetty, 2011).

A few households were involved in production of biofortified food crops in the study area or even aware of existence of such food crops. Major reason for this could be lack of assured market for these crops as reported by Waized *et al*. (2015) that farmers prefer to produce crops that can be used for both selling and consumption. Uncertainty of the market of the produce discourages the farmers from risking their scarce resources to engage in production (Bouis *et al*. 2013). Both the local agricultural and nutrition officers admitted that knowledge about these crops, including their health benefits, were quite low among the community members and therefore their demand was minimal. Also, prices for their seeds can also be a stumbling block, as majority of the rural farmers are small holder farmers with limited resources (Thierfelder *et al*., 2013). It was reported by one ward extension officer that “*few people that are aware of the benefits of biofortified food crops fail to grow them because they are required to purchase the seeds, which are relatively expensive due to low economic level of the majority of people in the area”*.

#### (iv) Fish farming

According to AUC-NEPAD (2014), more than 30% of Africans eat fish as their primary source of animal protein, making fish a good source of protein, micronutrients, and important fatty acids for addressing a variety of nutritional issues (Obiero et al. 2019). Over the past few years, fish farming has expanded, and more than 40% of the fish consumed worldwide is now a result. Beveridge et al. (2013) and De Graaf & Garibaldi (2014) reported that despite this expansion, the technique is still in its infancy in Sub-Saharan nations like Tanzania, where it is only sometimes used.

Study results show that, only a small number of farmers in the region engaged in fish farming. Mwima et al. (2012) reported the same in other Tanzanian regions. This observation may be due, in part, to the study subjects’ lack of knowledge regarding fish farming techniques. For individuals to appropriate execute the practice, they must be properly trained and support services from experts are also required. A study conducted by Mwajiande and Lugendo (2015) across 8 regions of Tanzania’s mainland, including Morogoro, the lack of extension services to assist farmers with managing their fish farming operations was reported as a significant obstacle to the adoption and practice of fish farming. It has been shown that fish farming sectors in Africa progressively contribute to food and nutrition security, employment, and livelihood support services and can highly help to confront the burden of malnutrition by serving as both the livelihood and income source to household members. (De Graaf & Garibaldi, 2014; FAO, 2018).

### Awareness about Nutrition-sensitive agriculture

Agriculture can play a significant role in enhancing the nutrition status of the household through various pathways including being source of food, employment, income, women empowerment and providing easy access to nutrients rich foods (FAO, 2014).

The findings indicate that most household heads were aware of the benefit of agriculture for dietary needs, particularly as a significant source of employment and income. But increased nutrition is not always a direct result of agricultural revenue (Ruel and Alderman, 2013). According to Anderson and Corps (2012), farmers often sell the nutrient-dense foods they grow, including vegetables, and instead purchase less nutrient-dense staples like rice, beans, and sugar. According to the district agricultural and nutrition officers, as well as the ward and village executive officers, the situation in the study region was similar in that farmers do sell their farm products to make money for obtaining other food items. The study also revealed that some respondents were not aware of important aspects of benefits of agriculture to nutrition such as enabling dietary diversity, allowing gender equity and flexibility of women to care for children in the household. One reason for this might be low nutrition knowledge, as more than two thirds of the respondents reported to have not received any nutrition education or training. Robinson *et al*. (2014) reported that most of the rural individuals who are normally poor farmers have low nutrition awareness especially on nutrient contents of the foods they produce or available in their areas. FAO (2017) stressed on the importance of providing nutrition education to the farmers because many causes of poor nutrition are engrained in their eating behavior such as negative attitudes towards fruits and vegetables, agricultural production decisions, intra-household food distribution in the households and child feeding practices that can be influenced by education.

Also, majority of them were not practicing other important ways of nutrition-sensitive agriculture apart from home gardening and small livestock keeping. However, the latter was being carried out not with a primary goal of enhancing nutritious household diet but to gain income. A study by Bundala *et al*. (2019) in Morogoro and Dodoma regions reported similar findings that producing animals for household consumption was not usually prioritized due to other desires, with income earning being a top priority. Other reason for this observation might include lack of adequate extension services for both agriculture and nutrition to provide awareness and knowledge to the farmers as it was cemented by Jaenicke and Virchow (2013). This was emphasized by the District nutrition officer who claimed that, the availability of only one nutrition specialist in the district does not correspond to the needs of the people in all villages in the district.

## Conclusion

The study assessed ways and awareness about nutrition-sensitive agriculture among farmers in Morogoro Rural District. Majority of the households practiced home gardening while a number of them kept livestock and these were reported to be conducted as normal practices in the community rather than being influenced by experts such as agriculture, livestock and nutrition officers. A very few households practiced other ways of nutrition-sensitive agriculture such as watering in production of fruits and vegetables, fish farming, sustaining production of nutrient-dense foods through the use of soil fertility conserving techniques, and minimize the use of industrial fertilizer through the use of farm yard manure. Economic hardship among most of the rural poor farmers can be one of the reasons that have led to the observed low practices of other ways of nutrition - sensitive agriculture especially those involving costs such as purchasing seeds of the improved food crops including biofortified ones. Lack of effective agriculture, livestock and nutrition extension services can also be blamed for the observed deficiencies. The study argues that there is a need for the government and partners to subsidize seeds and other agro-inputs and establish assured local market chain for nutrient-dense foods. On the other hand, key stakeholders and government should work together to ensure that community is educated on the benefit of producing and consuming nutrient dense foods.

## Data Availability

All data produced in the present study are available upon reasonable request to the authors

## Acknowledgement

Authors wish to acknowledge the Department of Nutrition and Consumer Sciences for their support during the preparation of this manuscript; Morogoro district council administration and farmers from Mkuyuni and Mvuha wards for providing data that made this study possible.

